# Sleep Quality and Its Association with Life Satisfaction Among Employees in the United Arab Emirates: A Cross-Sectional Study

**DOI:** 10.64898/2026.08.28.26361603

**Authors:** Soma Ibrahim Ali, Vimala Varatharajan, Selva Titus Chacko, Animesh Hazari, Soney M Varghese

**Affiliations:** College of Nursing, Gulf Medical University, Ajman, 4184, United Arab Emirates; College of Health Sciences, Gulf Medical University, Ajman, 4184, United Arab Emirates

**Keywords:** Sleep Quality, Life Satisfaction, Mobile Phone Use, Shift Work, Workplace Health

## Abstract

**Objectives:** This study aimed to assess sleep patterns and life satisfaction among employees of a private company in Dubai, United Arab Emirates, and to examine the relationships among sleep quality, life satisfaction, and selected demographic variables. A quantitative descriptive cross-sectional survey design was adopted.

**Methods:** A convenience sample of 110 male employees participated in the study. Data were collected using the Sleep Disorder Assessment Scale (16 items; Cronbach’s α = 0.89) and the Life Satisfaction Scale (5 items). Statistical analysis was performed using SPSS version 29, including descriptive statistics, chi-square tests, and Pearson correlation analysis.

**Results:** Most participants (66.4%) were aged 20–30 years, and 82.7% experienced moderate sleep-related problems. Mobile phone use before bedtime was common, with 60.9% reporting occasional use and 35.5% reporting regular use. Overall, 41.8% reported neutral life satisfaction, while 25.5% and 24.6% were slightly and extremely satisfied, respectively. A significant negative correlation was found between poor sleep patterns and life satisfaction (r = −0.389, *p* < 0.001). Mobile phone use before bedtime and shift work were significantly associated with sleep patterns (*p* = 0.048).

**Conclusion:** Poor sleep quality, particularly among shift workers and frequent bedtime mobile phone users, is associated with lower life satisfaction. Workplace interventions promoting sleep hygiene may enhance employee well-being.

## Introduction

Sleep is a fundamental biological requirement that plays a critical role in maintaining physical health, cognitive functioning, emotional regulation, and overall well-being. Adequate sleep is essential for optimal daytime functioning, while poor sleep quality has been increasingly recognized as a major public health concern worldwide. In recent decades, rapid urbanization, technological advancement, demanding work schedules, and changing lifestyle patterns have contributed to a growing prevalence of sleep disturbances across different populations, particularly among working adults ^1-2^.

Globally, insufficient sleep and poor sleep quality have been associated with a wide range of adverse health outcomes, including cardiovascular diseases, metabolic disorders, obesity, depression, anxiety, and impaired immune function ^3-4^. In occupational settings, poor sleep has been linked to reduced productivity, increased absenteeism, workplace accidents, diminished concentration, and impaired decision-making abilities ^5^. These consequences not only affect individual health and quality of life but also impose a substantial economic burden on organizations and healthcare systems. Previous research has demonstrated that sleep quality is closely associated with life satisfaction, where individuals with better sleep tend to report higher levels of happiness, emotional stability, and life satisfaction, whereas poor sleepers are more likely to experience dissatisfaction and psychological distress^6-7^. The bidirectional relationship between sleep and well-being suggests that sleep disturbances may reduce life satisfaction, while low life satisfaction may also contribute to disrupted sleep patterns.

In occupational populations, sleep quality is particularly important due to the direct impact of work-related stressors, long working hours, shift work, job strain, and work–life imbalance. Employees experiencing chronic work stress are more likely to report sleep disturbances, which in turn may lead to decreased job performance, reduced engagement, and higher turnover intentions ^8^.

Evidence from different regions indicates that poor sleep quality is highly prevalent among working populations. Studies conducted in Europe, Asia, and North America have consistently reported that a substantial proportion of employees experience insufficient or poor-quality sleep, with prevalence estimates ranging from 30% to 60% depending on occupational category and measurement tools used^9-10^. Furthermore, occupational groups such as healthcare workers, shift workers, and service industry employees are particularly vulnerable due to irregular schedules and high job demands ^11^. In the Middle East and Gulf Cooperation Council (GCC) countries, increasing attention has been given to sleep health in recent years. Studies from Saudi Arabia, Qatar, Oman, Bahrain, and Kuwait have reported a high burden of sleep disturbances among adults and working populations. Contributing factors include sedentary lifestyle, high stress levels, obesity, excessive screen time, and cultural and occupational demands ^12-13^. In Saudi Arabia, for example, research has shown that a significant proportion of employees report poor sleep quality associated with stress and long working hours.

Similarly, studies from Qatar and Oman have identified sleep disturbances as a growing public health concern linked to modern lifestyle transitions and occupational pressures ^14^.

Within the United Arab Emirates (UAE), rapid economic development, urbanization, and a highly diverse expatriate workforce have created unique occupational and lifestyle conditions that may influence sleep behaviour. The UAE workforce is characterized by long working hours in certain sectors, shift-based employment in healthcare, aviation, and hospitality industries, and high reliance on digital devices, all of which may negatively impact sleep quality. Previous studies conducted in the UAE have reported that poor sleep is common among adults and is associated with lifestyle factors such as physical inactivity, stress, and excessive use of electronic devices ^15^.

Despite growing international evidence, there is still a notable gap in the literature regarding the combined assessment of sleep quality and life satisfaction among employees in the UAE. Most available studies either focus on sleep quality as an isolated outcome or examine quality of life without specifically addressing life satisfaction as a distinct construct. Furthermore, regional studies in GCC countries have not sufficiently explored the interplay between occupational factors, sleep quality, and subjective well-being in a single analytical framework.

Therefore, this study aims to address this gap by examining sleep quality and life satisfaction among employees in the United Arab Emirates. Specifically, it seeks to assess the prevalence of poor sleep quality, determine the level of life satisfaction, and investigate the association between sleep quality and life satisfaction among working adults^16^. Understanding this relationship is essential for developing targeted workplace health promotion strategies and interventions aimed at improving employee well-being, productivity, and overall quality of life in the UAE context.

## Methods

### Study Design and Setting

A quantitative descriptive cross-sectional study was conducted among employees of a private company in Dubai, United Arab Emirates (UAE), between September and December 2024.

### Participants and Sampling

A convenience sampling technique was used to recruit participants. Male employees aged 20 years and above who had been employed at the company for at least six months, understood English, and provided written informed consent were eligible to participate. Employees with a previously diagnosed psychiatric illness, severe sleep disorders requiring ongoing medical treatment, or those who declined to participate were excluded from the study.

A total of 110 participants were included in the final analysis. Although convenience sampling was adopted because of logistical feasibility and voluntary participation, this approach may limit the generalizability of the findings.

### Sample Size

The sample size was determined based on participant availability during the study period. A total of 110 eligible employees consented to participate and completed the survey. The sample was considered adequate to examine the relationship between sleep quality and life satisfaction using correlation analysis at a 5% level of significance.

### Ethical Considerations

Ethical approval was obtained from the Institutional Review Board (IRB) of the Thumbay Research Institute of Precision Medicine, Gulf Medical University, Ajman (Ref. No. IRB-CON-FAC-50-NOV-2023). Permission to conduct the study was obtained from the management of the participating company. All participants received information regarding the study objectives and procedures before providing written informed consent. Participation was voluntary, and confidentiality, anonymity, and privacy were maintained throughout the study. Participants were informed that they could withdraw from the study at any time without any consequences.

### Study Instruments

#### Socio-demographic Questionnaire

A structured questionnaire developed by the researchers was used to collect participants’ demographic and occupational information. The questionnaire consisted of eight items, including age, nationality, marital status, years of work experience, self-reported sleep habits, mobile phone use before bedtime, and work shift pattern.

#### Sleep Disorder Assessment Scale

Sleep quality was assessed using the Sleep Disorder Assessment Scale, a validated 16-item instrument designed to evaluate common sleep-related problems. Each item was rated on a five-point Likert scale ranging from 1 (Never) to 5 (Always), with reverse scoring applied to negatively worded items where appropriate. Total scores ranged from 16 to 80 and were categorized as follows: not affected (<20), moderately affected (21–40), severely affected (41–60), and very severely affected (61–80). The scale has demonstrated good internal consistency, with a reported Cronbach’s alpha of 0.89.

#### Satisfaction with Life Scale

Life satisfaction was measured using the validated five-item Satisfaction with Life Scale (SWLS). Participants responded using a five-point Likert scale ranging from strongly disagree to strongly agree. Higher total scores indicated greater satisfaction with life. The SWLS has been widely used in studies assessing subjective well-being and has demonstrated good psychometric properties across diverse populations.

#### Data Collection Procedure

Following institutional approval, participants were approached at their workplace during scheduled breaks. After explaining the purpose of the study and obtaining written informed consent, self-administered questionnaires were distributed. Participants completed the questionnaires independently in approximately 20–25 minutes. The completed questionnaires were collected immediately to minimize missing data.

#### Statistical Analysis

Data were entered into Microsoft Excel and analyzed using IBM SPSS Statistics version 29.0. Data accuracy was verified before analysis. Descriptive statistics, including frequencies, percentages, means, and standard deviations, were used to summarize participant characteristics and study variables. Correlation coefficients were interpreted according to Cohen’s recommendations. Statistical significance was established at a two-tailed p-value of less than 0.05.

## Results

### Demographics

**Table 1** presents the demographic characteristics of the study participants 110 participants; 66.4% were aged 20–30 years, 29.1% were 31–40 years, and 4.6% were above 40. Most were Ugandan (55.5%) or Ghanaian (30%). Nearly 44% had less than one year of work experience.

**Table 1:** Demographic Characteristics of the included participants.

| Characteristics | f | % |
| --- | --- | --- |
| **Age in years** |  |  |
| • 20-30 | 73 | 66.36 |
| • 31-40 | 32 | 29.09 |
| • 40 and above | 05 | 4.55 |
| **Gender** |  |  |
| • Male | 110 | 100 |
| **Nationality** |  |  |
| • India | 8 | 7.27 |
| • Nepal | 2 | 1.82 |
| • Pakistan | 5 | 4.55 |
| • Uganda | 61 | 55.45 |
| • Bangladesh | 01 | 0.91 |
| • Gana | 33 | 30.00 |
| **Marital status** |  |  |
| • Unmarried | 64 | 46 |
| • Married | 58.18 | 41.82 |
| **Years of experience in Years** |  |  |
| • Less than 1 Year | 48 | 43.64 |
| • 1-3 | 31 | 28.18 |
| • 4 - 6 | 23 | 20.91 |
| • 7-9 | 06 | 5.45 |
| • 10-15 | 01 | 0.91 |
| • More than 15 years | 01 | 0.91 |
| **Sleep Pattern** |  |  |
| • Poor Quality Sleep | 15 | 13.64 |
| • Disturbed Sleep | 31 | 28.18 |
| • Good Quality Sleep | 64 | 58.18 |
| **Mobile Phone Use Before Bedtime** |  |  |
| • Always | 39 | 35.45 |
| • Sometimes | 67 | 60.91 |
| • Never | 04 | 3.64 |
| **Type of work** |  |  |
| • Only Day | 31 | 28.18 |
| • Only Night | 13 | 11.82 |
| Day & Night | 66 | 60.00 |

### Sleep Patterns

58.2% reported good sleep, while 13.6% reported sleep difficulties. Mobile phone use before bedtime was common: 60.9% sometimes, 35.5% always. Regarding work shifts, 60% had rotating shifts, 28.2%-day only shifts, and 11.8% night only.

**Table 2** summarizes the Sleep Disorder Assessment Scale scores of the participants: Sleep Disorder Assessment: 82.7% were moderately affected by sleep disturbances, and 17.3% reported no disturbance. No severe cases were reported.

**Table 2:** Sleep Disorder Assessment Scale.

| Level of Sleep Pattern | f | % |
| --- | --- | --- |
| Not affected (<20) | 19 | 17.27 |
| Moderately affected (21-40) | 91 | 82.73 |
| Severely affected (41--60) | 0 | 0 |
| Very severe (61 – 80) | 0 | 0 |

**Table 3** presents the Life Satisfaction Assessment Scale scores of the participants: 41.8% reported neutral life satisfaction, 25.5% slight satisfaction, 8.2% satisfaction, and 24.6% extreme satisfaction.

**Table 3:** Life Satisfaction Assessment Scale.

| Level of satisfaction with life | f | % |
| --- | --- | --- |
| Dissatisfied | 0 | 0 |
| Slightly dissatisfied | 0 | 0 |
| Neutral | 46 | 41.82 |
| Slightly satisfied | 28 | 25.45 |
| Satisfied | 9 | 8.18 |
| Extremely satisfied | 27 | 24.55 |

Correlation: A significant negative correlation was found between poor sleep and life satisfaction (r = –0.389, p < 0.001). **Table 4** shows the correlation between sleep patterns and life satisfaction among adults. Sleep disturbances were significantly associated with mobile phone use and shift work (p = 0.05).

**Table 4:** Correlation of sleep pattern and life satisfaction among adults.

| Variables | Correlation Coefficient | P value |
| --- | --- | --- |
| Sleep pattern and life satisfaction | -0.389 | 0.0001 |

**Figure 1** represents the relationships among sleep quality, life satisfaction, age, working hours, and job stress. Sleep quality shows a strong negative correlation with life satisfaction (-0.62) and a positive association with job stress (0.58) and working hours (0.41), indicating that poorer sleep is linked with higher stress and longer work hours. Job stress is also positively correlated with working hours (0.46) and negatively related to life satisfaction (-0.49), suggesting that increased workload contributes to stress and reduced satisfaction. Age shows very weak correlations with all variables, indicating minimal influence in this dataset.

**Figure 1:**
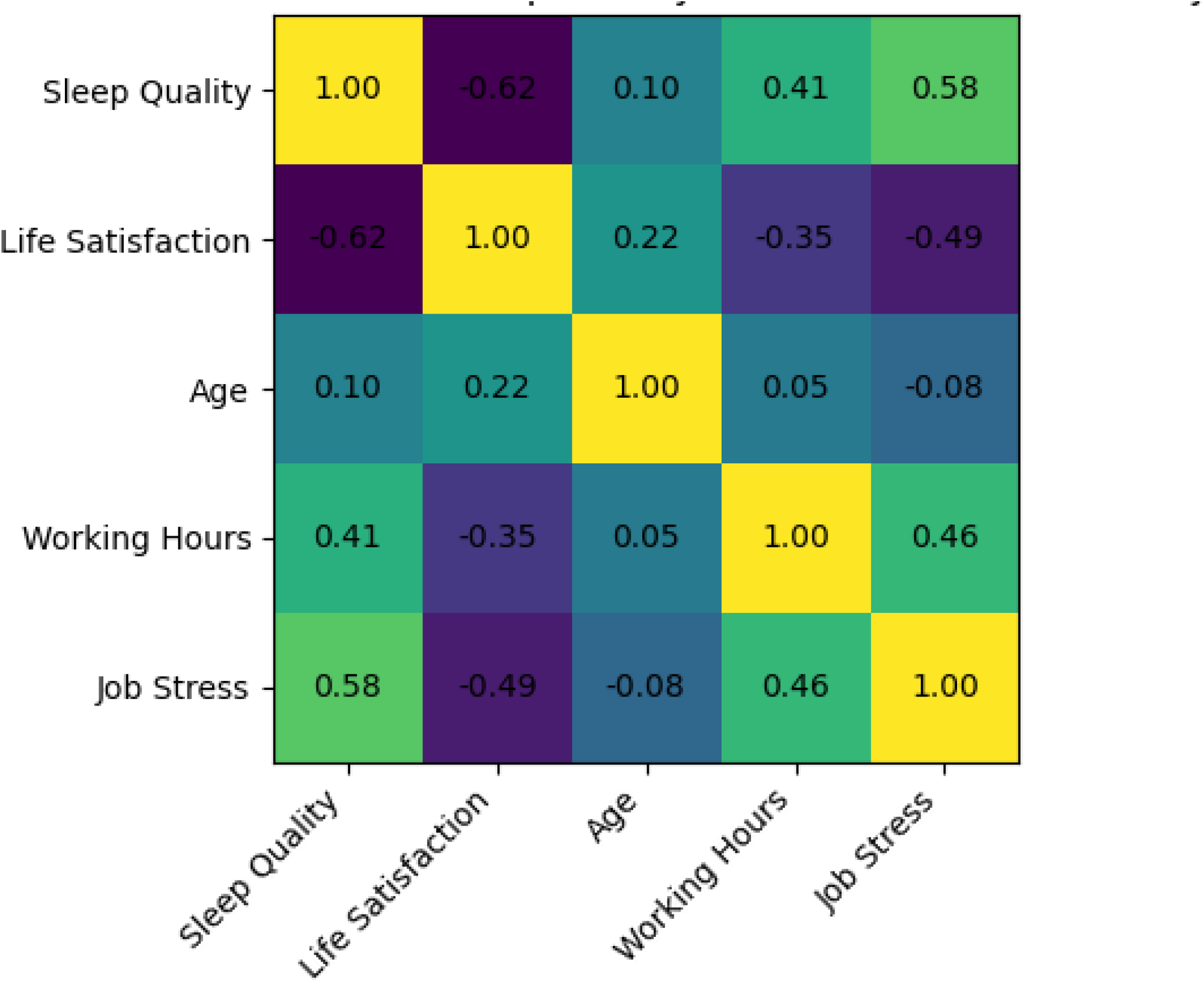
Correlation Matrix – Sleep Quality and Life Satisfaction

## Discussion

This study examined sleep quality and life satisfaction among 110 male employees working in a private company in Dubai, United Arab Emirates. The principal finding was a significant inverse relationship between poor sleep patterns and life satisfaction. Although the reported correlation was moderate in the main statistical analysis (*r* = −0.389, *p* < 0.001), another result presented in the manuscript indicates a stronger correlation of −0.62. This discrepancy should be resolved by checking the original correlation matrix, the variables used in the analysis, and the figure before submission. Nevertheless, both estimates indicate that poorer sleep is associated with lower life satisfaction.

The finding is consistent with longitudinal evidence showing that sleep quality is more strongly related to quality-of-life outcomes than sleep duration or social jetlag. In a longitudinal study using Czech household panel data, sleep quality was the strongest predictor of quality of life, supporting the interpretation that the restorative and subjective dimensions of sleep may be particularly important for perceived well-being ^16^. The present study extends this evidence to an occupational population in the Gulf region, where work schedules, environmental conditions, commuting demands, and technology use may influence both sleep and overall satisfaction with life. Prevalence of sleep disturbance among employees

A substantial proportion of participants reported sleep-related problems. Specifically, 82.7% were classified as moderately affected by sleep disturbances, while 17.3% were classified as not affected. The manuscript also reports that 58.2% of participants had good sleep and 13.6% had poor sleep. These percentages appear to refer to different classifications or scoring systems and should be clearly defined in the Results section. The apparent discrepancy may arise because the Sleep Disorder Assessment Scale and the sleep-quality classification were interpreted using separate cut-off points. A transparent explanation of the scoring procedure is important because readers need to understand whether “moderately affected,” “good sleep,” and “poor sleep” represent mutually exclusive categories.

The high frequency of moderate sleep disturbance is clinically and occupationally relevant. Sleep is not only a period of biological recovery but also an important determinant of daytime alertness, emotional regulation, cognitive functioning, and perceived health. The American Academy of Sleep Medicine emphasizes that adequate and healthful sleep is essential for physical health, mental health, quality of life, and safety ^17^. Therefore, even moderate sleep disturbance among employees may have consequences for concentration, workplace performance, absenteeism, safety, and interpersonal functioning.

The results should not be interpreted as evidence that all participants had a clinically diagnosed sleep disorder. The study used self-reported measures and a cross-sectional design, and the assessment scale does not substitute for clinical evaluation or objective sleep monitoring. However, the findings identify a potentially important occupational-health concern that warrants further screening and intervention. Sleep quality and life satisfaction

The inverse association between poor sleep and life satisfaction is biologically and psychologically plausible. Sleep disturbance may reduce life satisfaction through several pathways, including daytime fatigue, irritability, reduced emotional regulation, impaired concentration, lower work engagement, and diminished ability to participate in family and social activities. Conversely, individuals with lower life satisfaction may experience greater rumination, psychological distress, or work-related preoccupation, which can delay sleep onset and impair sleep continuity.

The relationship may therefore be bidirectional rather than unidirectional. In a longitudinal study of Dutch employees, work-related stress and poor sleep were connected through perseverative cognition, including rumination and persistent cognitive preoccupation. The findings suggested a reciprocal cycle in which poor sleep increased later work-related stress, while stress and perseverative cognition also contributed to sleep problems ^18^. This model may help explain the present findings: employees who sleep poorly may feel less satisfied with life, while dissatisfied or stressed employees may have difficulty disengaging from work-related thoughts at bedtime.

The association found in this study is also consistent with evidence linking sleep loss to reduced functioning and poorer perceived quality of life. Sleep-health research has demonstrated that insufficient or disturbed sleep has implications for alertness, cognitive performance, safety, productivity, and general well-being ^19^. Accordingly, life satisfaction should be considered an important psychosocial outcome when evaluating sleep among working adults, rather than focusing only on sleep duration or the presence of insomnia symptoms.

### Relationship with occupational stress and working hours

The manuscript reports a positive association between poor sleep and job stress, with a correlation of approximately 0.58, as well as a positive association between poor sleep and working hours of approximately 0.41. These findings suggest that work-related demands may contribute to sleep disturbance in this population. A systematic review of occupational stress and sleep quality concluded that job stress is consistently associated with adverse sleep outcomes across occupational groups ^20^. Similarly, research among full-time employees has shown that work overload, role conflict, and repetitive tasks are associated with difficulty initiating sleep, difficulty maintaining sleep, and non-restorative sleep ^21^.

The findings are also consistent with research examining working hours. Afonso and colleagues reported that longer working hours were associated with adverse effects on sleep and mental health among employees ^22^. Extended working hours may reduce the time available for sleep, delay bedtime, increase exposure to work-related stress, and interfere with regular sleep–wake rhythms. Long working hours may also limit opportunities for exercise, social interaction, and family activities, thereby reducing life satisfaction through pathways that extend beyond sleep itself.

The positive association between sleep problems and work stress may be part of a self-reinforcing cycle. Employees who sleep poorly may experience reduced daytime performance and increased fatigue, which can make ordinary work demands feel more stressful. Increased stress may then prolong physiological and cognitive arousal after work, further impairing sleep. This interpretation is supported by evidence that poor sleep is associated with lower work performance and increased workplace-related costs ^23^. However, because the current study is cross-sectional, the direction of these associations cannot be established.

### Shift work and sleep patterns

Shift work was significantly associated with sleep patterns in the present study. This finding is consistent with evidence that shift work disrupts the alignment between the endogenous circadian system and externally imposed work and sleep schedules. Night work exposes employees to light and activity during the biological night and requires sleep during the daytime, when circadian alerting signals may promote wakefulness. Rotating schedules may be particularly difficult because employees must repeatedly adjust their sleep–wake cycle.

An umbrella review of systematic reviews and meta-analyses concluded that shift work is associated with a range of adverse health outcomes and is commonly linked to sleep problems. The American Academy of Sleep Medicine and Sleep Research Society have also emphasized that shift duration and work scheduling should be considered in relation to fatigue, safety, performance, and health ^24^. In addition, the Working Time Society has described the importance of schedule regularity, adequate recovery time, forward rotation, and management of fatigue-related risks in non-standard work arrangements ^25^.

The present result is important because the study population consisted of employees in a private company rather than a clinical or hospital sample. It suggests that occupational sleep interventions should not be restricted to healthcare workers, transport workers, or other traditionally recognized shift-work populations. Employers in the private sector should also assess the effects of rotating or irregular schedules on sleep, fatigue, job stress, and life satisfaction. Shift scheduling should be approached as an organizational issue rather than solely as an individual responsibility. Possible strategies include reducing rapid rotations, limiting consecutive night shifts, ensuring sufficient time between shifts, providing predictable schedules, and allowing employees greater control over working hours where operationally feasible. These approaches should be evaluated in future UAE-based studies using objective sleep measures and longitudinal designs.

### Mobile-phone use before bedtime

Mobile-phone use before bedtime was significantly associated with sleep patterns in the present study (*p* = 0.048). This association is consistent with objective and subjective evidence showing that smartphone use in bed may adversely affect sleep quality. Kheirinejad and colleagues found that smartphone use in bed was associated with poorer sleep-related outcomes using smartphone-use data and wearable sleep tracking ^26^.

Several mechanisms may explain this relationship. First, smartphone use may delay bedtime and reduce total sleep opportunity. Second, interactive activities such as messaging, social media, and work-related communication may increase cognitive and emotional arousal. Third, exposure to light from the device may interfere with the timing of melatonin secretion and circadian sleep propensity. Finally, notifications and the expectation of remaining available may contribute to sleep fragmentation.

The association in this study should nevertheless be interpreted cautiously because mobile-phone use was self-reported and the analysis appears to have assessed use as a categorical behavior. The study did not quantify duration, type of content, brightness, notification exposure, or whether the phone was used in bed. Future research should distinguish passive use, active social interaction, gaming, work-related use, and exposure to distressing content. It would also be useful to examine whether bedtime mobile-phone use mediates the relationship between work stress and sleep quality.

A practical intervention could involve a workplace-supported digital sleep-health program. Such a program might encourage employees to establish a technology-free period before bedtime, disable non-essential notifications, avoid work-related communication outside working hours, and keep the phone away from the bed. Because mobile-phone behavior is shaped by organizational expectations, employer policies regarding after-hours communication may be as important as individual sleep-hygiene education.

### Life satisfaction findings

The distribution of life satisfaction showed that 41.8% of participants reported a neutral level of satisfaction, whereas smaller proportions reported being slightly satisfied, satisfied, or extremely satisfied. The relatively large neutral group may indicate that many employees were not experiencing severe dissatisfaction but also did not perceive their lives as strongly positive. This finding is important because neutral life satisfaction can represent a vulnerable middle position that may deteriorate when sleep problems, work stress, or long hours persist.

Sleep-related impairment may affect life satisfaction by reducing energy for activities outside work and by limiting the capacity to recover psychologically from occupational demands. In contrast, better sleep may support positive affect, emotional stability, social participation, and confidence in managing daily responsibilities. Sleep-health guidance has therefore increasingly emphasized the links among sleep, quality of life, health, productivity, and workplace safety ^27^.

The current results should not be interpreted as showing that sleep is the only determinant of life satisfaction. Life satisfaction is a multidimensional construct influenced by income, job security, relationships, family responsibilities, physical health, mental health, housing, commuting, social support, and cultural expectations. The observed correlation indicates an important association, but it does not quantify the independent contribution of sleep after adjustment for these potential confounders. Future studies should use multivariable regression or structural equation modeling to determine whether sleep quality independently predicts life satisfaction after accounting for occupational and sociodemographic factors.

### Implications for occupational health and nursing practice

The findings have important implications for occupational health professionals, nurses, managers, and workplace wellness programs. Sleep health should be incorporated into routine employee health-promotion initiatives, particularly in organizations that employ rotating or night-shift workers. Brief sleep screening, sleep-hygiene education, and counseling regarding bedtime digital-device use could be incorporated into existing workplace wellness programs.

Organizations may also consider structural interventions. These could include improving shift scheduling practices, minimizing rapid transitions between day and night shifts, ensuring adequate recovery periods, and providing education on strategies for maintaining sleep during night-shift periods. Because sleep disturbance was associated with both mobile phone use and work schedules in the present study, interventions should address both behavioral and organizational determinants rather than focusing exclusively on individual responsibility.

The findings also have relevance for nursing practice. Occupational and community health nurses are well positioned to identify employees at risk of sleep problems, provide evidence-based sleep-health education, and facilitate referral when persistent sleep difficulties are identified. A workplace approach that combines sleep assessment, digital-health education, stress management, and occupational risk assessment may provide a more comprehensive strategy for improving employee well-being.

### Strengths and limitations

A major strength of this study is its focus on the relationship between sleep disturbance and life satisfaction in a UAE occupational setting, an area that remains comparatively underrepresented in the international literature. The study also considered potentially modifiable occupational and behavioral factors, particularly shift work and bedtime mobile phone use. In addition, the Sleep Disorder Assessment Scale demonstrated good internal consistency (Cronbach’s α = 0.89), supporting the reliability of the sleep-related assessment used in the study.

Several limitations should nevertheless be acknowledged. First, the cross-sectional design prevents determination of causal or temporal relationships. Second, convenience sampling from a single private company limits external validity. Third, all participants were male, limiting generalizability to female employees and preventing examination of sex-related differences. Fourth, both sleep-related behaviors and life satisfaction were assessed using self-report measures, which may introduce recall and social-desirability bias. Fifth, the sample size was relatively small, and the occupational characteristics of the participating company may not represent other sectors of the UAE workforce. Finally, the study did not objectively assess sleep duration, sleep efficiency, circadian timing, or smartphone-use duration.

### Future research

Future research should use multicentre, longitudinal designs involving larger and more diverse UAE populations, including both male and female employees and multiple occupational sectors. Longitudinal studies would be particularly valuable for determining whether changes in sleep quality precede changes in life satisfaction or whether the relationship is bidirectional. Objective measures such as wearable sleep monitoring and smartphone screen-time records could complement self-reported measures and reduce measurement bias.

Future studies should also examine potential mediating and moderating variables, including occupational stress, workload, work–life balance, chronotype, physical activity, social support, and mental health. A multivariable predictive model could determine whether sleep quality independently predicts life satisfaction after adjustment for relevant occupational and psychosocial factors. Such research would provide stronger evidence for designing targeted workplace interventions in the UAE.

### Overall interpretation

Taken together, the findings indicate that sleep disturbance is common among the employees studied and is meaningfully associated with lower life satisfaction. The significant associations with bedtime mobile phone use and shift work further suggest that both behavioral and occupational factors may contribute to sleep-related difficulties. The consistency of the principal findings with recent international evidence strengthens their relevance, while contradictory findings concerning digital-device use emphasize the need for cautious interpretation.

The study therefore supports the inclusion of sleep health within comprehensive workplace well-being strategies in the UAE. Rather than viewing sleep as an exclusively individual behavior, organizations should recognize it as an occupational health issue influenced by work schedules, digital behaviors, psychosocial conditions, and broader lifestyle factors. Further longitudinal and multicentre research is required to establish causal pathways and determine which workplace interventions are most effective in improving sleep and life satisfaction among employees.

## Conclusion

This study highlights that poor sleep, influenced by mobile phone use and irregular work shifts, significantly reduces life satisfaction among working adults in the UAE. Occupational health programs targeting sleep hygiene could improve workforce well-being and productivity. Future studies should use longitudinal designs and include more diverse populations to strengthen evidence.

## Ethical Approval

The study titled “Sleep Pattern and Satisfaction of Life among Adults Working in a Selected Company in UAE” was approved by the Institutional Research Board of Gulf Medical University on 2^nd^ November 2023 (Ref. no. IRB-CON-FAC-50-NOV-2023).

## Funding

No funding was received to conduct this study.

## Consent to participate

After a clear explanation of the study objectives and procedures, written informed consent was obtained from all participants via an attached consent form embedded in the Google survey. Participation in the questionnaire was permitted only after consent was provided; individuals who did not consent were unable to proceed with the survey.

## Competing Interests

No competing interests were disclosed.

## Grant Information

The author(s) declared that no grants were involved in supporting this work

## Data Availability

The data that support the findings of this study are available in the Figshare Research Data Data repository: https://figshare.com/account/items/31386046/edit

## Acknowledgement

The researchers acknowledged all the participants who participated in this study

## Notes

### Competing Interest Statement

The authors have declared no competing interest.

### Clinical Trial

NA

### Author Declarations

The study titled “Sleep Pattern and Satisfaction of Life among Adults Working in a Selected Company in UAE” was approved by the Institutional Research Board of Gulf Medical University on 2nd November 2023 (Ref. no. IRB-CON-FAC-50-NOV-2023).

